# Serum PFAS and Lipid Concentrations in Ecuadorian Adolescents

**DOI:** 10.1101/2024.10.21.24315635

**Authors:** Michelle Guerra, Georgia Kayser, Harvey Checkoway, Jose Suarez-Torres, Dolores Lopez, Danilo Martinez, Carin A. Huset, Lisa A. Peterson, Jose Ricardo Suarez-Lopez

## Abstract

There is growing evidence that per- and polyfluoroalkyl substances (PFAS) may alter serum lipid concentrations; however, this topic is understudied in adolescents and Latin American populations. We aimed to characterize these associations among adolescents in Ecuador’s main floricultural region.

This cross-sectional study included 97 adolescents ages 11-17 years from Pedro Moncayo County, Ecuador. Generalized estimating equation models were applied to estimate the associations of serum perfluorooctane sulfonic acid (PFOS), perfluorooctanoic acid (PFOA), and perfluorononanoic acid (PFNA) concentrations with serum lipids. Models were adjusted for age, gender, height, body mass index (BMI), acetylcholinesterase (AChE) activity, and hemoglobin concentrations.

We observed statistically significant sex-specific associations of all PFAS with triglycerides. Significant inverse relationships between PFAS and triglycerides were observed in females (% lipid difference per 50% increase in: PFOS= −15.0% [95%CI: −24.72, −4.06], PFNA= −25.49% [−36.93, −12.00], and PFOA= −16.55% [−28.16, −3.07]) but not in males. No associations were observed between total cholesterol, high-density lipoprotein (HDL), or low-density lipoprotein (LDL) cholesterol and any PFAS.

PFOS, PFOA, and PFNA were inversely associated with triglycerides in adolescent females but not males. Further characterization of sex-specific associations of PFAS and blood lipids in adolescents is warranted.

## 1. Introduction

Per- and polyfluoroalkyl substances (PFAS) are persistent pollutants that have contaminated large areas of the world. PFAS can be found in non-stick coatings, water-resistant textiles, firefighting foam, personal care products and in many other products. PFAS have also been detected in insecticides, including sumithrin, piperonyl butoxide, malathion, spiromesifen, and imidacloprid (Brusseau et al., 2020; Lasee et al., 2022; Sargent, 2020), where they are applied to prolong the duration of their effectiveness (Swedish Chemicals Agency, 2015; Z. Wang et al., 2017). Furthermore, PFAS-based pesticides, such as sulfluramid, can also contaminate agricultural soils (Costello & Lee, 2020; Nascimento et al., 2018). Their persistence in the environment, mobility, and bioaccumulation in agricultural settings is of concern. Of the greater than 15,000 types of PFAS that exist (National Institute of Environmental Health Sciences, 2023), some of the most prevalent in humans include perfluorooctane sulfonic acid (PFOS), perfluorooctanoic acid (PFOA), perfluorononanoic acid (PFNA), and perfluorohexane sulfonic acid (PFHxS) (Agency for Toxic Substances and Disease Registry, 2024)

Several epidemiologic studies have found associations between PFAS concentrations and increased risk of dyslipidemia (Centers for Disease Control and Prevention, 2024b; Ho et al., 2022). A 10-year longitudinal study of 864 Swedish participants, aged 70 years, indicated that PFOA, PFNA, and PFOS concentrations in serum were directly associated with triglycerides, low-density lipoprotein (LDL), and high-density lipoprotein (HDL) cholesterol (Dunder et al., 2022). Other cross-sectional studies in Italy and the US have demonstrated an increasing trend between increased triglyceride and LDL levels with PFOA, PFOS, and PFNA in serum concentrations (Canova et al., 2020; Batzella et al., 2022; Starling et al., 2019). In three studies that separated data by gender, found associations between PFOS, PFOA, and PFNA with higher LDL cholesterol levels in males. Conversely, direct associations were found between various PFAS and higher HDL cholesterol levels in females (Batzella et al., 2022; Canova et al., 2020; Starling et al., 2019). Contrasting with these findings, one study in Nunavik, Canada found an inverse association between PFOS and triglycerides in adult females, whereas an Italian study found no association (Château-Degat et al., 2010; Jain & Ducatman, 2019).

Few studies have examined the relations between PFAS and blood lipids among adolescents. In a three-year surveillance study on Italian adolescents, researchers found a significant direct association of PFOS and PFNA concentrations with total cholesterol, LDL and HDL levels, whereas PFOA and PFHxS were significantly inversely associated with HDL (Canova et al., 2021). Similar results were found in a large cross-sectional study conducted in Norway, of adolescents (Averina et al., 2021). Few studies of adolescents examined sex and age as effect modifiers for PFAS-blood lipid associations; those that do exist, show mixed results. In a large cross-sectional study of Italian adolescents, elevated PFOA and PFOS significantly positively correlated with HDL concentrations in females, but not in males (Canova et al., 2021). Contrary to these findings, a large study of adolescents living with PFAS-contaminated drinking water in mid–Ohio River Valley, USA, found a statistically significant positive association of PFOA with total cholesterol and LDL in males and younger children, but a weaker significant trend in females. Furthermore, the relationship between PFOS and HDL was statistically significant for males, but no relationship was observed in females (Frisbee et al., 2010).

The overall objective of this cross-sectional study was to analyze the association of PFAS with blood lipids in adolescents being raised in farming communities in northern Ecuador. Given the trends observed in studies of adults, we hypothesized that serum concentrations of PFOS, PFOA, and PFNA will have direct association with total cholesterol, HDL, LDL, and triglyceride concentrations. To the best of our knowledge, this is the first study to characterize the relationships between biomarkers of PFAS exposure and blood lipids in adolescents residing in agricultural areas and the first in a Latin American population.

## 2. Methods

### 2.1 Study Design and Recruitment

The Study of Secondary Exposures to Pesticides among Children and Adolescents (ESPINA) is a prospective cohort study designed to examine the health and developmental effects of exposures to agrochemicals on children residing in Pedro Moncayo County, Pichincha Province, Ecuador, known for its significant floricultural industry and other agricultural production (e.g. produce, fruits and grains).

The ESPINA study conducted participant examinations in 2008 and 2016. In 2008, an examination was conducted on 313 boys and girls, aged 4–9 years, residing in the agricultural County of Pedro Moncayo, Pichincha province, Ecuador, during July and August. In 2016, a follow-up examination included 554 participants aged 12–17 years, consisting of 238 from the 2008 study and 316 new volunteers. Of these 554 participants, 535 were examined between July and October 2016, while 331 were examined in April 2016, with 311 participants undergoing examinations in both April and July–October.

Recruitment in 2008 primarily relied on the 2004 Survey of Access and Demand of Health Services in Pedro Moncayo County, covering 71% of the County’s population. Fundación Cimas del Ecuador carried out this survey in partnership with the Governments of Rural Parishes of Pedro Moncayo and community members. Additional participants for the ESPINA study were recruited via community announcements, word-of-mouth, and a strategy that ensured balanced representation across different ages and sexes. Eligibility criteria included: A) having lived with a flower plantation worker for a minimum of one year; or B) never having lived with an agricultural worker, never having resided in a home where agricultural pesticides were stored, and never having had prior contact with pesticides. Further details about the methodologies used in the 2008 and 2016 studies have been previously published. (Suarez-Lopez et al., 2012; Chronister et al., 2023). In 2016, new participants were identified through the System of Local and Community Information (SILC) created by Fundación Cimas del Ecuador, using data from the 2016 Pedro Moncayo County Community Survey (previously the Survey of Access and Demand of Health Services in Pedro Moncayo County). As in 2008, all adolescent participants in 2016 reported not currently working in agriculture.

The present analyses include data from an ancillary study in which we measured serum lipids in 101 participants who were examined during the July-October 2016 period. Participants were selected into the pilot study based on: A) having been examined in the April 2016 examination; and B) having the highest and lowest levels of acetylcholinesterase activity, to include participants with potential for having both elevated and low organophosphate pesticide exposures in 2016 (Phillips et al., 2021). Four participants were excluded due to missing PFAS measurement; therefore, the current analysis includes data of 97 participants examined in July-October 2016. Figure 1 shows the flow chart of participants selected for the present analysis.

**Figure 1:**
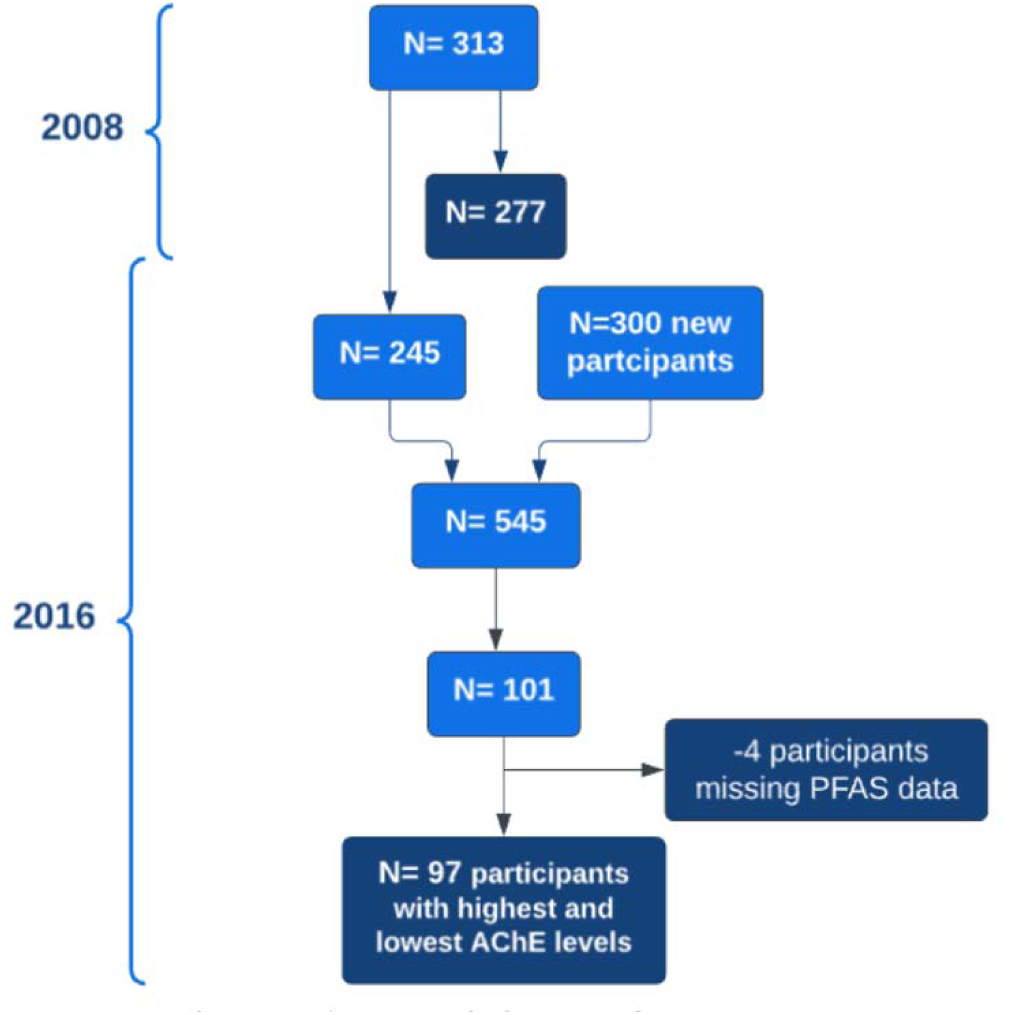
Participant flow chart.

This study received approval from the institutional review boards at the University of California, San Diego in the United States, and Universidad San Francisco de Quito in Ecuador. Additionally, the Ecuadorian Ministry of Health approved the study. For participants under 18 years of age, parents provided informed consent and assent for their children to participate. All aspects of this research were completed in accordance with the Helsinki Declaration.

### 2.2 Data Collection

In the July-October 2016 examination, pesticide exposure history, socioeconomic status, and demographic characteristics were collected through at-home interviews of parents. Participant examinations took place in 7 schools in Pedro Moncayo County. Participants’ weight was assessed using a digital scale (Tanita model 0108MC; Tanita Corporation of America, Arlington Heights, IL, USA), and their height was measured in millimeters using a height board, with individuals not wearing shoes or headwear during measurements. The World Health Organization (WHO) growth standards were used to calculate height-for age and BMI-for-age Z-scores (*WHO Multicentre Growth Reference Study*, 2006). Participants were not asked to fast prior to their examinations. Additional details of the examination methods have been described previously (Chronister et al., 2023).

### 2.3 PFAS Quantification in Serum

Blood samples were collected via venipuncture of the median cubital or cephalic veins of the arm. Samples were processed on site, and extracted serum was then aliquoted and frozen on dry-ice in coolers until they were transported to Quito, Ecuador (Netlab) for storage at −70°C. Samples were then shipped frozen (∼-15°C) using a specialized courier from Quito to the University of California San Diego (UCSD), San Diego, California, USA for long-term storage at −80°C. Samples remained frozen during transportation. Samples were then shipped frozen (∼- 15°C) from UCSD to the Minnesota CHEAR Exposure Assessment Hub.

In this study, the following PFAS were quantified: PFOS, PFOA, PFNA, PFOSA, PFHXS, and ETFOSA. However, the analysis of serum PFAS included compounds (PFOS, PFNA, and PFOA) that were detectable in at least 30% of participants. The analysis was performed through protein precipitation followed by centrifugation, concentration, and analysis using liquid chromatography tandem mass spectrometry (LC/MS/MS). In brief, a solution containing stable isotope-labeled internal standards and acetonitrile was added to a 400 µL serum sample in microcentrifuge tubes. After mixing and centrifugation, the resulting liquid was transferred to 96-well plates and concentrated using nitrogen. Following concentration, the sample underwent analysis using LC/MS/MS (Shimadzu Prominence coupled to AB Sciex 5500Q) with electrospray ionization and multiple reaction monitoring (MRM) within specified time frames. Calibration curves were prepared using bovine calf serum, and quantification was conducted using isotope dilution. Each analyte was monitored using at least 2 MRM transitions, and unknown samples were confirmed by matching retention times and ion ratios with the calibration curve. Furthermore, each batch of 20 unknown samples included a minimum of 3 quality control (QC) samples, consisting of a method blank, and low and high concentration levels. The quality control materials were prepared in bovine calf serum. An extra QC was performed if an unknown sample needed dilution, either because of insufficient sample volume or an initial concentration exceeding the calibration curve limits. Batches with QC recovery rates outside the 70-130% range were reanalyzed; in cases where a QC failure could not be resolved through reinjection/reanalysis, the resulting unknown data was qualified to reflect this result.

### 2.4 Acetylcholinesterase activity quantification

EQM Test-mate ChE Cholinesterase Test System 400 (EQM AChE Erythrocyte Cholinesterase Assay Kit 470; EQM Research, Inc, Cincinnati, OH) was used to measure erythrocytic AChE activity and hemoglobin concentrations, from a fresh finger-stick blood sample. Blood samples were, upon sample collection, analyzed at ambient temperatures between 15 and 28°C, which are within the recommended range, following standard procedures (EQM Research Inc., 2003).

### 2.5 Serum Lipid Concentrations

Total cholesterol, HDL-Cholesterol (HDL), LDL-Cholesterol (LDL), and triglycerides in serum were measured using enzymatic methods (OSR6616, OSR6695, OSR6196, and OSR66118 kit; Beckman Coulter, Brea USA) using a Beckman Coulter AU680 Chemistry Analyzer at US Specialty Labs in San Diego, CA (Beckman Coulter, n.d.).

### 2.6 Statistical Analysis

The mean or prevalence of characteristics was calculated for all participants and stratified by tertiles of PFOS concentration. P-trend values for participant characteristics were computed through a generalized linear model (GLM), using a log-transformed continuous variable for PFOS concentration. The geometric means of serum PFAS concentrations were computed both for the entire sample size and categorized by gender.

To estimate associations between PFAS and blood lipids we included PFAS that were detected in at least 30% of participants. PFOS and PFOA were detected in all participants, and PFNA was detected in 43.3%. For values below the level of quantitation (LOQ), we imputed concentrations by calculating the LOQ/√2. The associations between PFAS and blood lipids were calculated using generalized estimating equations (GEE). Models adjusted for age, sex, z-score for height-for-age, z-score for BMI-for-age, AChE activity, and hemoglobin concentration. Models adjusted for AChE activity, a physiological marker of exposure to cholinesterase inhibitor pesticides, considering that participants lived in agricultural settings and greater exposure to pesticides that may contain PFAS (Sargent, 2020; Brusseau et al., 2020; Lasee et al., 2022) may be associated with greater serum PFAS concentrations, and AChE activity may be negatively associated with blood lipids. and hemoglobin concentration (Pothu et al., 2019). Hemoglobin concentration was adjusted for in the models to correct AChE activity values considering that we measured erythrocytic AChE activity (EQM Research Inc., 2003).

Statistical significance was determined at an alpha level of 0.05. We used natural log transformation on both the independent and dependent variables since they were not normally distributed. We converted the beta coefficient to reflect the percent difference in serum lipid concentrations per a 50% increase in PFAS concentration by raising 1.5 to the power of the beta coefficient, subtracting one, and then multiplying by 100.

We assessed the effect modification by age and gender using multiplicative interaction terms. Associations with significant gender or age interaction (P<0.05) were then stratified by a categorical variable of the effect modifier. SAS 9.4 was used for all analyses in this study.

## 3. Results

### 3.1 Participant Characteristics

Of the 97 participants, 54% were males and 46% were females, and had a mean age of 14.7 years. The mean z-score for height-for-age was −1.4 SD, whereas the mean z-score for BMI-for-age was 0.3 SD. Males had higher concentrations of PFOS than females, and borderline positive associations were observed between AChE activity and PFOS concentration (**Table 1**). Compared to the full cohort, prior to excluding participants with unmeasured serum lipids, (n=523 with PFAS measures), the present subset of participants had comparable (minimally higher) concentrations of PFOS (0.57 ng/mL vs 0.54 ng/mL), PFOA (0.33 ng/mL vs 0.32 ng/mL) and PFNA (0.093 ng/mL vs 0.088 ng/mL).

**Table 1:**
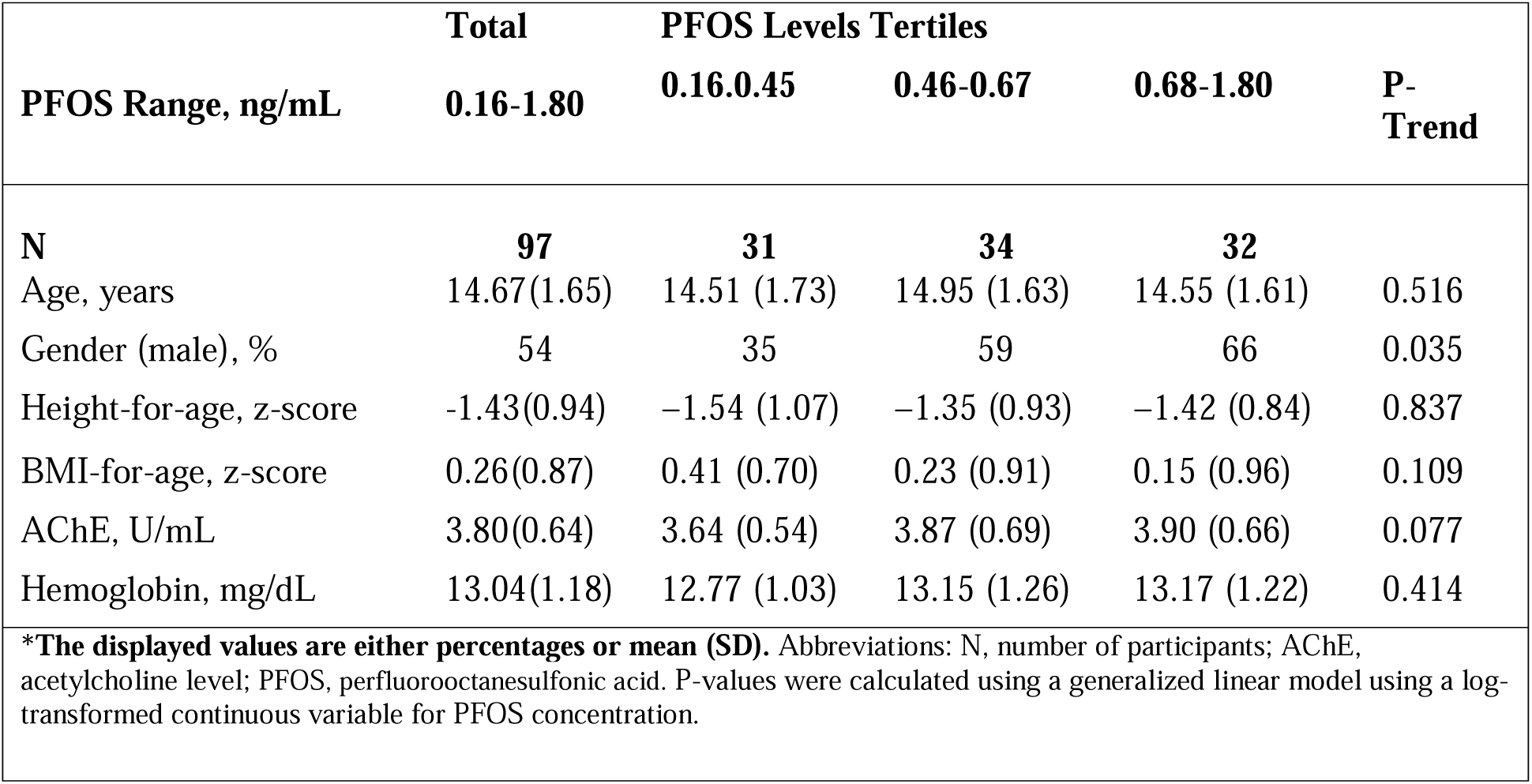
Participant characteristics by tertiles of PFOS.

**Table 2:**
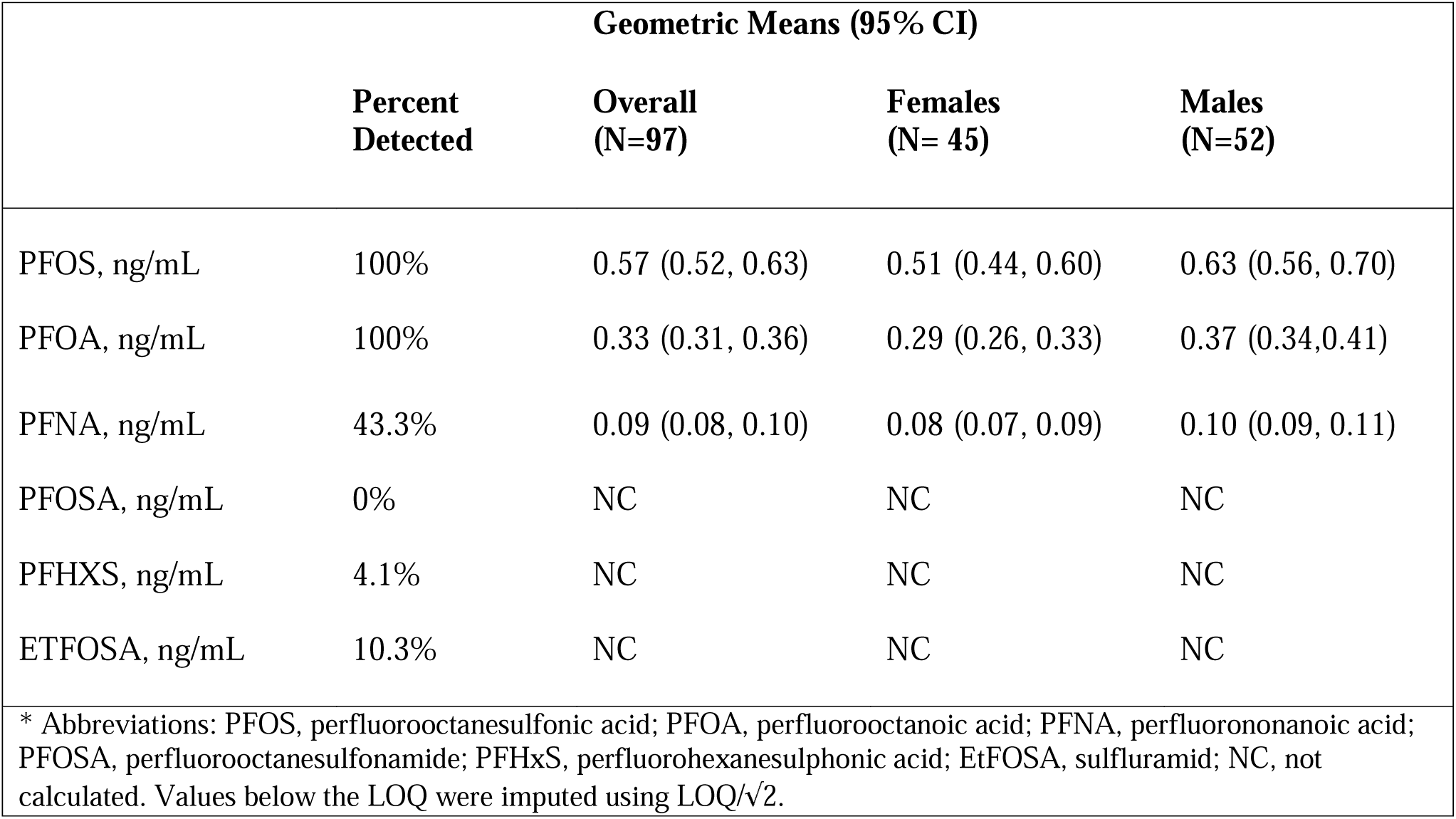
Concentrations of serum PFAS in adolescent participants of the ESPINA study.

### 3.2 PFAS and Serum Lipids

We did not observe any associations between PFOA, PFOS or PFNA with total cholesterol, HDL, or LDL. However, we detected a statistically significant negative association between PFOA and triglycerides, with triglycerides decreasing by 10.86% (95% CI: −20.14 to - 0.51) for every 50% increase in PFOA (ng/mL, **Table 3**). Negative, albeit non-significant associations were observed with PFOS and PFNA. Our analyses showed evidence of effect modification by gender in the associations of triglycerides with PFOS (p=0.02), PFOA (p=0.04) and PFNA (p<0.001). In stratified analyses, we found a statistically significant relationship between PFOS, PFOA, and PFNA with triglycerides in females but not in males (**Table 4**). In females, PFNA exhibited the strongest inverse association with triglycerides (for every 50% increase in PFNA, triglycerides were lower by 25.5% (95% CI: −36.9% to −12.0%), followed by PFOA (16.6%, 95% CI: −28.2% to −3.10%) and PFOS (−15% (95% CI: −36.9% to −4.10%). No evidence of curvilinear associations was observed in any of these analyses.

**Table 3:**
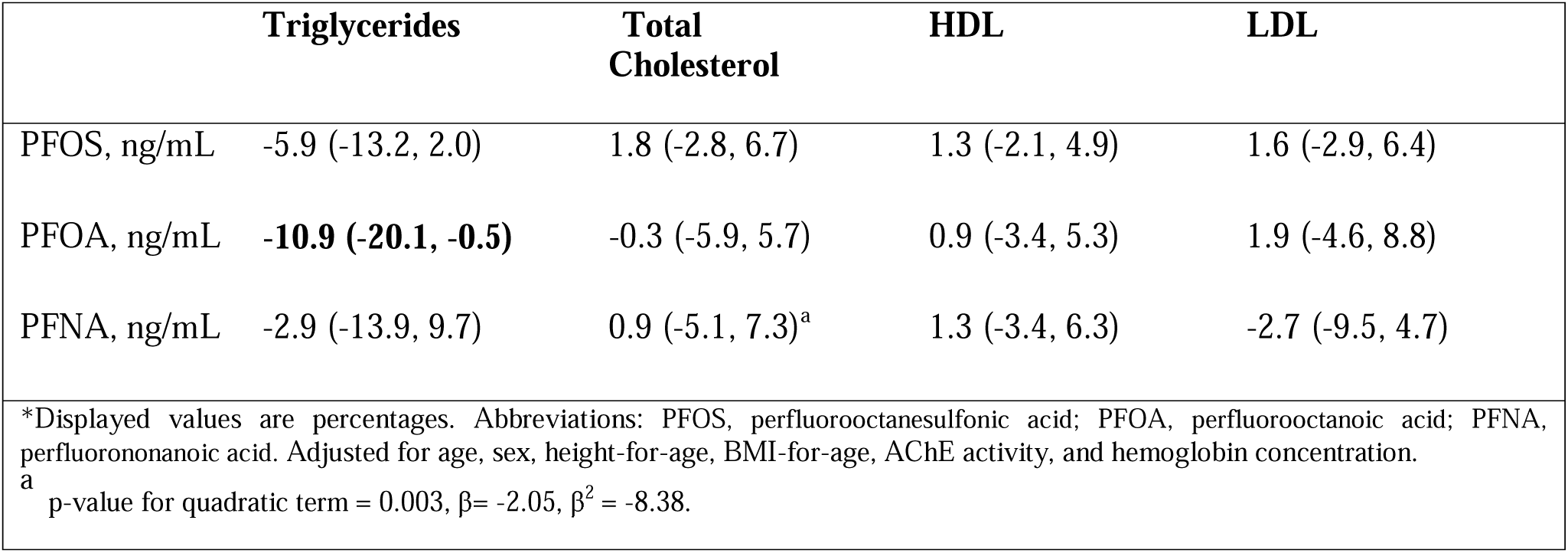
Percent difference of blood lipids per 50% increase in PFAS concentration (95% CI).

**Table 4:**
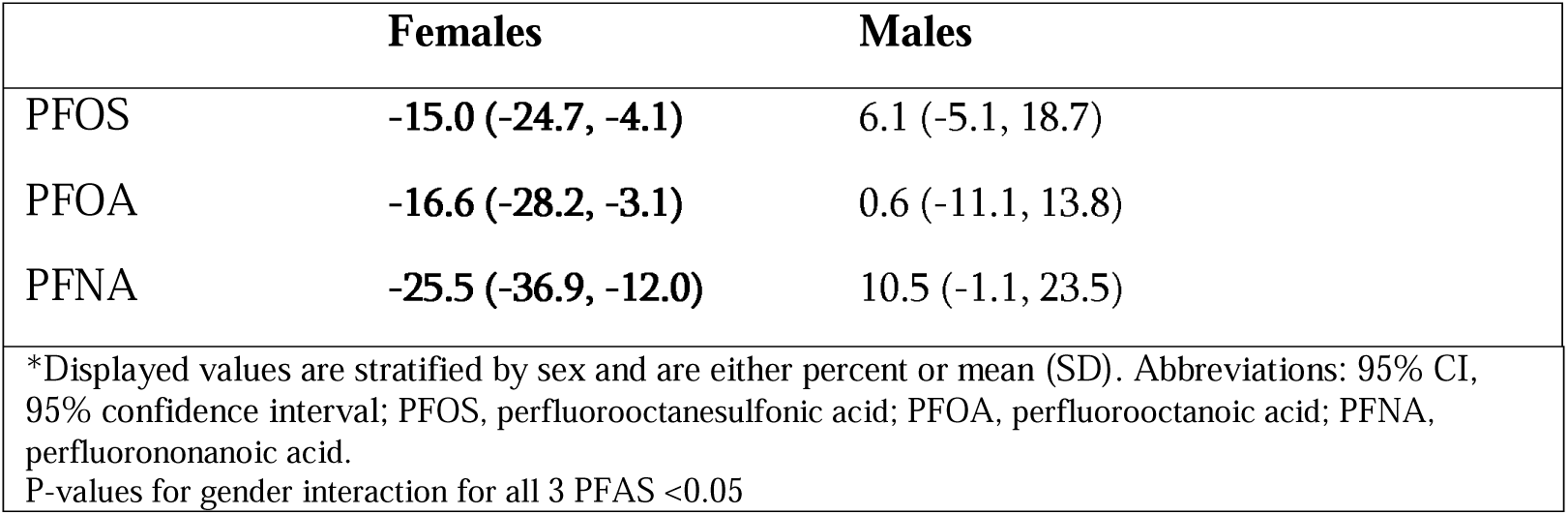
Percent difference in triglyceride concentration (mg/dL) per 50% increase in PFAS concentration (95% CI), stratified by sex.

## 4. Discussion

Our findings revealed a significant inverse association of PFOS, PFNA, and PFOA with triglyceride concentrations in female adolescents, but not in males. We also found no associations of PFOA, PFNA, or PFOS with total cholesterol, HDL, or LDL. Few adolescent studies have analyzed the relationship between PFAS and triglyceride concentrations in sex-stratified models and none have been conducted in populations in agricultural settings or in Latin America. The concentrations of PFAS in our study population were nearly six times lower for PFOS (3.35 µg/L vs 0.57 µg/L) and PFNA (0.54 µg/L vs 0.09 µg/L), and about five times lower for PFOA (1.5 µg/L vs 0.3 µg/L) compared to adolescents (12-19 years) examined in the National Health and Nutrition Examination Survey (NHANES) 2011-2018 in the USA (Centers for Disease Control and Prevention, 2024a).

Two studies of adolescents have evaluated effect modification by sex on the association between PFAS and triglycerides. A cross-sectional study of 12,476 children and adolescents in Mid–Ohio River Valley, USA found similar results to our findings, reporting that PFOS were inversely associated with triglycerides in females but not males. Furthermore, researchers found that PFOA serum concentrations were directly associated with increased LDL and total cholesterol (Frisbee et al., 2010). However, in a cross-sectional study in the USA involving 877 adolescents, no association was found when the analysis adjusted for sex and age, despite not specifically assessing modification by sex (Geiger et al., 2014). In a study on adults living in northern Canada, PFOS and PFHxS concentrations were also inversely associated with triglycerides only in females (Château-Degat et al., 2010). Whereas in obese and non-obese participants in the USA, PFOS and PFHxS were also positively associated with triglycerides among both males and females (Jain & Ducatman, 2019).

Contrary to our findings, most adolescent studies have reported direct associations of PFOA and PFOS with total cholesterol and LDL in sex-adjusted models (Geiger et al., 2014; Frisbee et al., 2010; Averina et al., 2021; Koshy et al., 2017; Zeng et al., 2015). However, our findings are consistent with some studies in adolescents (Kang et al., 2018) and adults (Predieri et al., 2015), that did not observe associations of PFOS, PFOA, and PFNA with total cholesterol, HDL, or LDL.

The observed inverse associations between serum PFAS and triglycerides among females in this study suggest a sex-specific hypolipidemic effect of PFAS. While lower triglycerides are generally beneficial, it is crucial to consider their potential impact on essential organ functions during adolescence, where critically low triglyceride levels can adversely affect growth and development (Miller et al., 2011; National Research Council (US) Committee on Diet and Health, 1989; Zhao et al., 2021).

PFAS may alter triglyceride concentrations in the human body by disrupting hepatic lipid metabolism (Tan et al., 2013; Jain & Ducatman, 2019; Arvind et al., 2000). Furthermore, PFAS may influence lipid regulation-associated genes, such as the microsomal triglyceride transfer protein (MTTP) gene (Marques et al., 2022), or affect the expression of the lipoprotein lipase (LPL) gene responsible for producing LPL, which breaks down triglycerides from lipoproteins (H. Wang & Eckel, 2009). Further research is needed to determine how triglyceride concentrations may affect PFAS.

The reason for the observed effect modification by sex in this study is unclear and further research is needed to understand it. Sex-specific variations in PFAS concentrations may be influenced by physiological factors unique to females, such as menstruation and lactation, which could facilitate more efficient elimination of PFAS from the body (Kingsley et al., 2018; Lorber et al., 2015; Wong et al., 2014). Furthermore, the differential impact of PFAS on lipoproteins in females compared to males could be attributed to the interaction of certain PFAS compounds like PFOS and PFOA with estrogen receptors, which are more prevalent in females (Mokra, 2021). This interaction could disrupt estrogen signaling pathways, potentially affecting lipid metabolism, and contributing to adverse health outcomes. Sex-specific hormonal and genetic differences play pivotal roles in lipoprotein metabolism (Palmisano et al., 2018). Estrogen, for instance, has been shown to enhance lipoprotein metabolism by increasing levels of HDL (high-density lipoprotein), which aids in removing cholesterol from the bloodstream (Lamon-Fava et al., 2006). These hormonal differences suggest that females may exhibit more efficient triglyceride metabolism compared to males. Additionally, sex-related genetic variations in lipid transport, metabolism, and synthesis can influence the composition and distribution of lipoproteins in the blood (Paththinige et al., 2017). Therefore, a comprehensive investigation into sex-based distinctions in liver enzymes, hepatic lipid metabolism, and cholesterol biomarkers is warranted to elucidate the observed effect modification by sex in the association between PFAS exposure and serum lipids.

Limitations of this study include a small sample size, which limits the statistical power to properly characterize the associations of PFAS with blood lipids. Despite this, we were able to detect strong associations between PFAS and triglycerides, even in analyses stratified by gender. It is plausible that additional associations of PFAS with other blood lipid components will be detected if we were to include a larger subset of ESPINA study participants. Another limitation is the cross-sectional study design which precludes us from understanding the long-term effects of PFAS exposure and the directionality of the association. Further research assessing effect modification by sex and potential mechanisms for this sex difference is needed to better understand PFAS exposure and serum lipids in adolescents as research is nascent. Moreover, conducting repeat-measures study is warranted. Future research should also assess the cellular and molecular mechanisms responsible for the effects of PFAS on lipoproteins to provide insight into the physiological impact of PFAS on lipoprotein metabolism.

## 5. Conclusion

In this study, we found that the association between serum PFAS levels and triglycerides was significantly modified by sex. Specifically, PFOS, PFOA, and PFNA were inversely associated with triglyceride levels in female adolescents but not in male adolescents. Additionally, we did not observe associations of PFAS with other blood lipids. This study expands the limited research available on the prevalence of PFAS in serum in rural Latin American populations and their relationships with serum lipids. We highlight the need for more research in these populations to increase our understanding of sex-specific relationships between PFAS and serum lipids.

## Data Availability

All data produced in the present study are available upon reasonable request to the authors

## 6. Acknowledgments

Research reported in this publication was supported by the National Institute of Environmental Health Sciences of the National Institutes of Health under Award Numbers (R01ES025792, R01ES030378, R21ES026084, U2CES026533). NIEHS funding was also provided to Dr. Georgia Kayser in grant numbers K01ES031697. We thank ESPINA study staff, Fundación Cimas del Ecuador, the Parish Governments of Pedro Moncayo County, community members of Pedro Moncayo and the Education District of Pichincha-Cayambe-Pedro Moncayo counties for their contributions and support on this project. We thank Karin Vevang and Kitrina M. Barry for their assistance in quantifying PFAS in our biospecimens. We also thank Dr. Rajendra Parajuli for edits on this manuscript.

## References

Agency for Toxic Substances and Disease Registry. (2024, January 18). Blood testing for PFAS. https://www.atsdr.cdc.gov/pfas/health-effects/blood-testing.html

Arvind, A., Osganian, S. A., Cohen, D. E., & Corey, K. E. (2000). Lipid and Lipoprotein Metabolism in Liver Disease. In K. R. Feingold, B. Anawalt, M. R. Blackman, A. Boyce, G. Chrousos, E. Corpas, W. W. de Herder, K. Dhatariya, K. Dungan, J. Hofland, S. Kalra, G. Kaltsas, N. Kapoor, C. Koch, P. Kopp, M. Korbonits, C. S. Kovacs, W. Kuohung, B. Laferrère, … D. P. Wilson (Eds.), Endotext. MDText.com, Inc. http://www.ncbi.nlm.nih.gov/books/NBK326742/

Averina, M., Brox, J., Huber, S., & Furberg, A.-S. (2021). Exposure to perfluoroalkyl substances (PFAS) and dyslipidemia, hypertension and obesity in adolescents. The Fit Futures study. Environmental Research, 195, 110740. 10.1016/j.envres.2021.110740

Batzella, E., Zare Jeddi, M., Pitter, G., Russo, F., Fletcher, T., & Canova, C. (2022). Associations between Mixture of Perfluoroalkyl Substances and Lipid Profile in a Highly Exposed Adult Community in the Veneto Region. International Journal of Environmental Research and Public Health, 19(19), Article 19. 10.3390/ijerph191912421

Beckman Coulter. (n.d.). AU680 Clinical Chemistry Analyzer. Retrieved May 20, 2024, from https://www.beckmancoulter.com/products/chemistry/au680

Brusseau, M. L., Anderson, R. H., & Guo, B. (2020). PFAS concentrations in soils: Background levels versus contaminated sites. Science of The Total Environment, 740, 140017. 10.1016/j.scitotenv.2020.140017

Canova, C., Barbieri, G., Zare Jeddi, M., Gion, M., Fabricio, A., Daprà, F., Russo, F., Fletcher, T., & Pitter, G. (2020). Associations between perfluoroalkyl substances and lipid profile in a highly exposed young adult population in the Veneto Region. Environment International, 145, 106117. 10.1016/j.envint.2020.106117

Canova, C., Di Nisio, A., Barbieri, G., Russo, F., Fletcher, T., Batzella, E., Dalla Zuanna, T., & Pitter, G. (2021). PFAS Concentrations and Cardiometabolic Traits in Highly Exposed Children and Adolescents. International Journal of Environmental Research and Public Health, 18(24), Article 24. 10.3390/ijerph182412881

Centers for Disease Control and Prevention. (2024a). Biomonitoring Data Tables for Environmental Chemicals. https://www.cdc.gov/exposurereport/data_tables.html?NER_SectionItem=NHANEShttps://www.cdc.gov/biomonitoring/PFOA_FactSheet.htm

Centers for Disease Control and Prevention. (2024b, February 27). LDL and HDL Cholesterol and Triglycerides. Cholesterol. https://www.cdc.gov/cholesterol/about/ldl-and-hdl-cholesterol-and-triglycerides.html

Château-Degat, M.-L., Pereg, D., Dallaire, R., Ayotte, P., Dery, S., & Dewailly, É. (2010). Effects of perfluorooctanesulfonate exposure on plasma lipid levels in the Inuit population of Nunavik (Northern Quebec). Environmental Research, 110(7), 710–717. 10.1016/j.envres.2010.07.003

Chronister, B. N. C., Yang, K., Yang, A. R., Lin, T., Tu, X. M., Lopez-Paredes, D., Checkoway, H., Suarez-Torres, J., Gahagan, S., Martinez, D., Barr, D., Moore, R. C., & Suarez-Lopez, J. R. (2023). Urinary Glyphosate, 2,4-D and DEET Biomarkers in Relation to Neurobehavioral Performance in Ecuadorian Adolescents in the ESPINA Cohort. Environmental Health Perspectives, 131(10), 107007. 10.1289/EHP11383

Costello, M. C. S., & Lee, L. S. (2020). Sources, Fate, and Plant Uptake in Agricultural Systems of Per- and Polyfluoroalkyl Substances. Current Pollution Reports. 10.1007/s40726-020-00168-y

Dunder, L., Lind, P. M., Salihovic, S., Stubleski, J., Kärrman, A., & Lind, L. (2022). Changes in plasma levels of per- and polyfluoroalkyl substances (PFAS) are associated with changes in plasma lipids—A longitudinal study over 10 years. Environmental Research, 211, 112903. 10.1016/j.envres.2022.112903

EQM Research Inc. (2003). Test-mate ChE Cholinesterase Test System (Model 400) Instruction … Yumpu.Com. https://www.yumpu.com/en/document/view/10197244/test-mate-che-cholinesterase-test-system-model-400-instruction-

Frisbee, S. J., Shankar, A., Knox, S. S., Steenland, K., Savitz, D. A., Fletcher, T., & Ducatman, A. M. (2010). Perfluorooctanoic Acid, Perfluorooctanesulfonate, and Serum Lipids in Children and Adolescents: Results From the C8 Health Project. Archives of Pediatrics & Adolescent Medicine, 164(9), 860–869. 10.1001/archpediatrics.2010.163

Geiger, S. D., Xiao, J., Ducatman, A., Frisbee, S., Innes, K., & Shankar, A. (2014). The association between PFOA, PFOS and serum lipid levels in adolescents. Chemosphere, 98, 78–83. 10.1016/j.chemosphere.2013.10.005

Ho, S. H., Soh, S. X. H., Wang, M. X., Ong, J., Seah, A., Wong, Y., Fang, Z., Sim, S., & Lim, J. T. (2022). Perfluoroalkyl substances and lipid concentrations in the blood: A systematic review of epidemiological studies. Science of The Total Environment, 850, 158036. 10.1016/j.scitotenv.2022.158036

Jain, R. B., & Ducatman, A. (2019). Roles of gender and obesity in defining correlations between perfluoroalkyl substances and lipid/lipoproteins. Science of The Total Environment, 653, 74–81. 10.1016/j.scitotenv.2018.10.362

Kang, H., Lee, H.-K., Moon, H.-B., Kim, S., Lee, J., Ha, M., Hong, S., Kim, S., & Choi, K. (2018). Perfluoroalkyl acids in serum of Korean children: Occurrences, related sources, and associated health outcomes. Science of The Total Environment, 645, 958–965. 10.1016/j.scitotenv.2018.07.177

Kingsley, S. L., Eliot, M. N., Kelsey, K. T., Calafat, A. M., Ehrlich, S., Lanphear, B. P., Chen, A., & Braun, J. M. (2018). Variability and predictors of serum perfluoroalkyl substance concentrations during pregnancy and early childhood. Environmental Research, 165, 247–257. 10.1016/j.envres.2018.04.033

Koshy, T. T., Attina, T. M., Ghassabian, A., Gilbert, J., Burdine, L. K., Marmor, M., Honda, M., Chu, D. B., Han, X., Shao, Y., Kannan, K., Urbina, E. M., & Trasande, L. (2017). Serum perfluoroalkyl substances and cardiometabolic consequences in adolescents exposed to the World Trade Center disaster and a matched comparison group. Environment International, 109, 128–135. 10.1016/j.envint.2017.08.003

Lamon-Fava, S., Postfai, B., Diffenderfer, M., DeLuca, C., O’Connor, J., Welty, F. K., Dolnikowski, G. G., Barrett, P. H. R., & Schaefer, E. J. (2006). Role of the Estrogen and Progestin in Hormonal Replacement Therapy on Apolipoprotein A-I Kinetics in Postmenopausal Women. Arteriosclerosis, Thrombosis, and Vascular Biology, 26(2), 385–391. 10.1161/01.ATV.0000199248.53590.e1

Lasee, S., McDermett, K., Kumar, N., Guelfo, J., Payton, P., Yang, Z., & Anderson, T. A. (2022). Targeted analysis and Total Oxidizable Precursor assay of several insecticides for PFAS. Journal of Hazardous Materials Letters, 3, 100067. 10.1016/j.hazl.2022.100067

Lorber, M., Eaglesham, G. E., Hobson, P., Toms, L.-M. L., Mueller, J. F., & Thompson, J. S. (2015). The effect of ongoing blood loss on human serum concentrations of perfluorinated acids. Chemosphere, 118, 170–177. 10.1016/j.chemosphere.2014.07.093

Marques, E., Pfohl, M., Wei, W., Tarantola, G., Ford, L., Amaeze, O., Alesio, J., Ryu, S., Jia, X., Zhu, H., Bothun, G. D., & Slitt, A. (2022). Replacement per- and polyfluoroalkyl substances (PFAS) are potent modulators of lipogenic and drug metabolizing gene expression signatures in primary human hepatocytes. Toxicology and Applied Pharmacology, 442, 115991. 10.1016/j.taap.2022.115991

Miller, M., Stone, N. J., Ballantyne, C., Bittner, V., Criqui, M. H., Ginsberg, H. N., Goldberg, A. C., Howard, W. J., Jacobson, M. S., Kris-Etherton, P. M., Lennie, T. A., Levi, M., Mazzone, T., & Pennathur, S. (2011). Triglycerides and Cardiovascular Disease. Circulation, 123(20), 2292–2333. 10.1161/CIR.0b013e3182160726

Mokra, K. (2021). Endocrine Disruptor Potential of Short- and Long-Chain Perfluoroalkyl Substances (PFASs)—A Synthesis of Current Knowledge with Proposal of Molecular Mechanism. International Journal of Molecular Sciences, 22(4), Article 4. 10.3390/ijms22042148

Nascimento, R. A., Nunoo, D. B. O., Bizkarguenaga, E., Schultes, L., Zabaleta, I., Benskin, J. P., Spanó, S., & Leonel, J. (2018). Sulfluramid use in Brazilian agriculture: A source of per- and polyfluoroalkyl substances (PFASs) to the environment. Environmental Pollution, 242, 1436–1443. 10.1016/j.envpol.2018.07.122

National Institute of Environmental Health Sciences. (2023). Perfluoroalkyl and Polyfluoroalkyl Substances (PFAS). National Institute of Environmental Health Sciences. https://www.niehs.nih.gov/health/topics/agents/pfc

National Research Council (US) Committee on Diet and Health. (1989). Diet and Health: Implications for Reducing Chronic Disease Risk. National Academies Press (US). http://www.ncbi.nlm.nih.gov/books/NBK218743/

Palmisano, B. T., Zhu, L., Eckel, R. H., & Stafford, J. M. (2018). Sex differences in lipid and lipoprotein metabolism. Molecular Metabolism, 15, 45–55. 10.1016/j.molmet.2018.05.008

Paththinige, C., Sirisena, N., & Dissanayake, V. (2017). Genetic determinants of inherited susceptibility to hypercholesterolemia – a comprehensive literature review. Lipids in Health and Disease, 16(1), 103. 10.1186/s12944-017-0488-4

Phillips, S., Suarez-Torres, J., Checkoway, H., Lopez-Paredes, D., Gahagan, S., & Suarez-Lopez, J. R. (2021). Acetylcholinesterase activity and thyroid hormone levels in Ecuadorian adolescents living in agricultural settings where organophosphate pesticides are used. International Journal of Hygiene and Environmental Health, 233, 113691. 10.1016/j.ijheh.2021.113691

Pothu, U. K., Thammisetty, A. K., & Nelakuditi, L. K. (2019). Evaluation of cholinesterase and lipid profile levels in chronic pesticide exposed persons. LWW, 8(6), 2073–2078. 10.4103/jfmpc.jfmpc_239_19

Predieri, B., Iughetti, L., Guerranti, C., Bruzzi, P., Perra, G., & Focardi, S. E. (2015). High Levels of Perfluorooctane Sulfonate in Children at the Onset of Diabetes. International Journal of Endocrinology, 2015. 10.1155/2015/234358

Sargent, S. (2020, December 1). Aerially Sprayed Pesticide Contains PFAS. PEER.Org. https://peer.org/aerially-sprayed-pesticide-contains-pfas/

Starling, A., Higgins, C. P., Barton, K., McDonough, C., Calafat, A., & Adgate, J. (2019). Exposure to per- and polyfluoroalkyl substances and cross-sectional associations with serum lipids in a highly exposed Colorado (US) population. LWW, 3, 382. 10.1097/01.EE9.0000610252.31693.5f

Suarez-Lopez, J. R., Jacobs, D. R., Himes, J. H., Alexander, B. H., Lazovich, D., & Gunnar, M. (2012). Lower acetylcholinesterase activity among children living with flower plantation workers. Environmental Research, 114, 53–59. 10.1016/j.envres.2012.01.007

Swedish Chemicals Agency. (2015). Occurrence and use of highly fluorinated substances and alternatives, Report 7/15.

Tan, X., Xie, G., Sun, X., Li, Q., Zhong, W., Qiao, P., Sun, X., Jia, W., & Zhou, Z. (2013). High Fat Diet Feeding Exaggerates Perfluorooctanoic Acid-Induced Liver Injury in Mice via Modulating Multiple Metabolic Pathways. PLOS ONE, 8(4), e61409. 10.1371/journal.pone.0061409

Wang, H., & Eckel, R. H. (2009). Lipoprotein lipase: From gene to obesity. American Journal of Physiology-Endocrinology and Metabolism. 10.1152/ajpendo.90920.2008

Wang, Z., DeWitt, J. C., Higgins, C. P., & Cousins, I. T. (2017). A Never-Ending Story of Per- and Polyfluoroalkyl Substances (PFASs)? Environmental Science & Technology, 51(5), 2508–2518. 10.1021/acs.est.6b04806

WHO Multicentre Growth Reference Study. (2006). https://www.who.int/tools/child-growth-standards/who-multicentre-growth-reference-study

Wong, F., MacLeod, M., Mueller, J. F., & Cousins, I. T. (2014). Enhanced Elimination of Perfluorooctane Sulfonic Acid by Menstruating Women: Evidence from Population-Based Pharmacokinetic Modeling. Environmental Science & Technology, 48(15), 8807– 8814. 10.1021/es500796y

Zeng, X.-W., Qian, Z., Emo, B., Vaughn, M., Bao, J., Qin, X.-D., Zhu, Y., Li, J., Lee, Y. L., & Dong, G.-H. (2015). Association of polyfluoroalkyl chemical exposure with serum lipids in children. Science of The Total Environment, 512–513, 364–370. 10.1016/j.scitotenv.2015.01.042

Zhao, Q., Chu, Y., Pan, H., Zhang, M., & Ban, B. (2021). Association between triglyceride glucose index and peak growth hormone in children with short stature. Scientific Reports, 11(1), Article 1. 10.1038/s41598-021-81564-2

